# Intrahepatic transcriptomics differentiate advanced fibrosis and clinical outcomes in adults with the Fontan circulation

**DOI:** 10.1101/2023.06.05.23290997

**Authors:** Katia Bravo-Jaimes, Xiuju Wu, Leigh C Reardon, Gentian Lluri, Jeannette P Lin, Jeremy P Moore, Glen Van Arsdell, Reshma Biniwale, Ming-Sing Si, Bita V Naini, Robert Venick, Sammy Saab, Christopher L Wray, Reid Ponder, Carl Rosenthal, Alexandra Klomhaus, Kristina I Böstrom, Jamil A Aboulhosn, Fady M Kaldas

**Affiliations:** Department of Cardiovascular Diseases. Mayo Clinic Jacksonville Florida; Ahmanson/UCLA Adult Congenital Heart Disease Center. University of California, Los Angeles; Division of Cardiology. Department of Medicine. University of California, Los Angeles; Department of Pediatric Cardiology. University of California, Los Angeles Mattel Children’s Hospital; Department of Pathology and Lab Services. University of California, Los Angeles; Department of Gastroenterology, Hepatology and Nutrition. University of California, Los Angeles Mattel Children’s Hospital; Department of Anesthesiology. University of California, Los Angeles; Dumont-UCLA Liver Transplant Center. Department of Surgery. University of California, Los Angeles; Department of Medicine Statistics Core. David Geffen School of Medicine. University of California, Los Angeles

**Keywords:** single ventricle, liver fibrosis, transcriptomics, Fontan-associated liver disease, Fontan outcomes

## Abstract

**Background:** The molecular mechanisms underlying Fontan associated liver disease (FALD) remain largely unknown. We aimed to assess intrahepatic transcriptomic differences among patients with FALD according to the degree of liver fibrosis and clinical outcomes.

**Methods:** This retrospective cohort study included adults with the Fontan circulation at the Ahmanson/UCLA Adult Congenital Heart Disease Center. Clinical, laboratory, imaging and hemodynamic data prior to the liver biopsy were extracted from medical records. Patients were classified into early (F1-F2) or advanced fibrosis (F3-F4). RNA was isolated from formalin-fixed paraffin embedded liver biopsy samples; RNA libraries were constructed using rRNA depletion method and sequencing was performed on Illumina Novaseq 6000. Differential gene expression and gene ontology analyses were carried out using DESeq2 and Metascape. Medical records were comprehensively reviewed for a composite clinical outcome which included decompensated cirrhosis, hepatocellular carcinoma, liver transplantation, protein-losing enteropathy, chronic kidney disease stage 4 or higher, or death.

**Results:** Patients with advanced fibrosis had higher serum BNP levels and Fontan, mean pulmonary artery and capillary wedge pressures. The composite clinical outcome was present in 23 patients (22%) and was predicted by age at Fontan, right ventricular morphology and presence of aortopulmonary collaterals on multivariable analysis. Samples with advanced fibrosis had 228 up-regulated genes compared to early fibrosis. Samples with the composite clinical outcome had 894 up-regulated genes compared to those without it. A total of 136 up-regulated genes were identified in both comparisons and these genes were enriched in cellular response to cytokine stimulus, response to oxidative stress, VEGFA-VEGFR2 signaling pathway, TGF-beta signaling pathway, and vasculature development.

**Conclusions:** Patients with FALD and advanced liver fibrosis or the composite clinical outcome exhibit up-regulated genes including pathways related to inflammation, congestion, and angiogenesis. This adds further insight into FALD pathophysiology.

---

Patients with a functional single ventricle represent a population with highly complex anatomy, physiology, and management challenges. Since its first description in 1971, the Fontan operation has been performed for palliation of single ventricle physiology^1^. Several evolutionary modifications have been made to improve long-term outcomes, but short-and long-term complications including Fontan-associated liver disease (FALD)^2^, continue to occur after Fontan palliation. Preliminary studies have demonstrated that hepatic fibrosis develops early after the Fontan palliation^3^ and will be present in virtually all patients^2–4^, depending mostly on duration of Fontan physiology rather than hemodynamics^4, 5^. Decompensated cirrhosis will only be apparent in some patients and a minority will develop hepatocellular carcinoma^5^. The pathophysiology of FALD includes chronic passive congestion due to elevated systemic venous pressures ^6, 7^, as well as chronic low cardiac output. This physiology results in decreased oxygen delivery to centrilobular cells, zone 3 hepatocytes atrophy, sinusoidal fibrosis, eventual bridging fibrosis and finally cardiac cirrhosis^8, 9^.

With >70,000 post-Fontan patients worldwide now reaching adulthood^2^, there is a need to better characterize the molecular pathways underlying FALD. We assessed intrahepatic gene expression profiles in adults with the Fontan circulation in comparison with donor controls to test the hypotheses that (1) despite clinical heterogeneity, patients with the Fontan circulation and advanced fibrosis exhibit a distinctive gene transcriptome that contrasts with that of early fibrosis and controls; (2) these differences involve enriched pathways related to angiogenesis; and (3) molecular phenotyping can identify Fontan subgroups that exhibit distinct clinical features and prognosis.

## METHODS

### Fontan study population

The Fontan patient group consisted of adults (>= 18 years old) who have had at least two visits at the Ahmanson/UCLA Adult Congenital Heart Disease Center between January 2005 and December 2021 and had tissue available from at least one liver biopsy. The study was approved by the UCLA Institutional Review Board. Baseline clinical, laboratory, imaging and hemodynamic information were extracted from medical records and the results closest to the date of liver biopsy within a 1-year period (6 months before and after the liver biopsy) were recorded. These variables included age, sex, height, weight, body mass index (BMI), race/ethnicity, cardiac anatomy, genetic syndromes, age at Fontan, cardiac medications (beta-blockers, antiplatelets, anticoagulants, loop diuretics, renin-angiotensin-aldosterone system inhibitors, phosphodiesterase 5 inhibitors, endothelin receptor antagonists, antiarrhythmics), diabetes, hypertension, obesity (BMI >30), hepatitis C, alcohol use, arrhythmias, NYHA class, single ventricle systolic function and atrioventricular valve regurgitation (echocardiographic or by cardiac magnetic resonance), creatinine, platelet count, hemoglobin, AST, ALT, GGT, INR, AFP, total bilirubin, BNP, liver ultrasound, MRI and CT characteristics, exercise-induced desaturation, peak VO2, VE/VCO2, high exercise capacity (>80% age, sex matched peak VO2)^10^, low exercise capacity (<50% age, sex matched peak VO2)^10^, Fontan pressures, pulmonary artery pressures, pulmonary capillary wedge pressures, single ventricle end-diastolic pressure, Qs, pulmonary vascular resistance (PVR), PVR index (PVRi), systemic vascular resistance (SVR), SVR index (SVRi), Fontan pathway obstruction (angiographic evidence of stenosis along the Fontan pathway with at least 1 mmHg gradient)^10^, diastolic dysfunction (single ventricle end-diastolic pressure or pulmonary capillary wedge pressure >= 12 mmHg at baseline or >= 15 mmHg after volume or contrast load)^10^, elevated PVR (PVRi >= 2 WU*m2)^10^, aortopulmonary and venous-venous collaterals (assessed by angiography).

### Liver tissue procurement and processing

Archived formalin-fixed paraffin embedded (FFPE) liver tissue from patients with the Fontan circulation who have had liver biopsies were processed by the Translational Pathology Core Laboratory. Slides were independently interpreted by two liver pathologists and METAVIR scores were determined by consensus. Advanced liver fibrosis was defined as those having METAVIR F3-F4 fibrosis on liver biopsy. Normal liver tissue and post-liver transplant biopsy tissue were used as controls.

### RNA preparation and sequencing

Five to six 10 µM sections from each FFPE tissue block were used for RNA extraction. Total RNA was isolated from the liver sections using RNeasy FFPE kit (Qiagen, Cat. No. 73504). RNA samples with DV200 ≥ 30% were processed for library construction using rRNA depletion methods (Roche, KAPA RNA HyperPrep Kit w/ Ribo Erase, KK8561). Sequencing was performed using Illumina Novaseq 6000 platform (2×100bp) to the depth of 28-110 million reads per library at UCLA Technology Center for Genomics & Bioinformatics. The FASTQ raw reads were mapped using Spliced Transcripts Alignment to a Reference (STAR, version 2.7.10a)^11^ to the human reference genome GRCh38 with default parameters. The counts for each gene were generated using –quantMode GeneCounts function along the STAR alignment.

### Differential gene expression analyses

Transcripts were first quality filtered to exclude the genes with a mean read count < 5 in each sample. Transcripts without gene annotation were also excluded using R package biomaRt. Data normalization and differential expression analyses were carried out using R package DESeq2 (version 1.38.1)^11^. After estimation of size factors and dispersion, the likelihood ratio test (LRT) with negative binomial generalized linear model (NB GLM) was used for fitting. Sequencing depth normalized gene counts were obtained from DESeqDataSet. Differentially expressed genes (DEGs) were defined using the 5% false discovery rate (Benjamini-Hochberg method) threshold and two-fold change for significance. To minimize the possible false discoveries, DESeq function were performed again following permutations on the variables of fibrosis or composite clinical outcome. The genes identified in the differential test after permutations were filtered out.

### Gene ontology and gene-disease association analyses

Gene ontology and gene-disease association (DisGeNet) enrichment analyses were performed using Metascape platform^12^. The DisGeNET platform integrates information of human gene-disease associations from various repositories^13, 14^.

### Statistical analysis of clinical variables

Continuous and categorical data were reported as median (interquartile range) and number (percentage), respectively. Between-group differences in the baseline variables described above were examined using Kruskal-Wallis or Pearson chi-square (or Fisher’s exact) tests as appropriate. Variables found to significantly (p<0.05) differ between the groups were used in covariate adjusted models. A composite clinical outcome included decompensated cirrhosis (ascites requiring paracentesis, esophageal variceal bleeding, or hepatic encephalopathy), hepatocellular carcinoma, need for liver transplantation, protein-losing enteropathy, chronic kidney disease stage 4 or higher, or death. An unadjusted cumulative incidence function and Gray’s Test, stratified by early vs advanced fibrosis was used to compare time to event relative to the date of Fontan between the two groups. A multivariable Cox proportional hazards model was built using covariates that were associated in univariate analysis as well as those considered clinically meaningful.

### Data availability

RNA sequencing raw data and normalized gene counts can be requested to the corresponding author upon reasonable request.

## RESULTS

### Baseline characteristics according to degree of fibrosis

Of 130 adults with the Fontan circulation who had 152 liver biopsies (20 patients had 2 liver biopsies and 2 had 3 liver biopsies), 106 patients with 112 adequate RNA quality samples were included. Fifty-five (51.8%) were women, 71 (66.9%) were white and 29 (27.4%) were Hispanic. Fifteen patients (14.2%) had no fibrosis, 40 (37.7%) had early fibrosis (F1 in 19 and F2 in 31) and 45 (48.1%) had advanced fibrosis (F3 in 31 and F4 in 14) (Supplemental figure 1). Baseline characteristics are detailed in Table 1. Though aortopulmonary collaterals were identified all groups, those without fibrosis were more likely to have aortopulmonary collaterals (46.7% without fibrosis, 10.0% with early fibrosis, 26.8% with advanced fibrosis, p=0.0332). Higher SVRi was seen in the group without fibrosis [25.7 WU·m2 (9.2) without fibrosis, 19.5 WU·m2 (11.3) with early fibrosis, 18.1 WU·m2 (9.4) with advanced fibrosis, p=0.014]. Those with advanced fibrosis were more likely to be on PDE-5 inhibitors (6.7% without fibrosis, 38.0% with early fibrosis, 43.9% with advanced fibrosis, p=0.033), have diastolic dysfunction (40.0% without fibrosis, 48.0% with early fibrosis, 70.7% with advanced fibrosis, p=0.017) and higher BNP [44 pg/mL (46) without fibrosis, 47 pg/mL (91) with early fibrosis, 95 pg/mL (130) with advanced fibrosis, p=0.016)]. Similarly, they also had a higher VE/VCO2 slope [(27.1 (6) without fibrosis, 27.7 (5.8) with early fibrosis, 30.9 (6.4) with advanced fibrosis, p=0.009], Fontan [14.0 mmHg (5.0) without fibrosis, 14.5 mmHg (3.0) with early fibrosis, 17.0 mmHg (6.0) with advanced fibrosis, p<0.001), mean pulmonary artery [13.5 mmHg (5.0) without fibrosis, 14.0 mmHg (3.0) with early fibrosis, 17.0 mmHg (5.0) with advanced fibrosis, p<0.001), and pulmonary capillary wedge [10.0 mmHg (4.0) without fibrosis, 10.0 mmHg (4.0) with early fibrosis, 12.0 mmHg (5.0) with advanced fibrosis, p=0.005) pressures.

**Table 1.** Baseline characteristics of adults with the Fontan circulation according to the degree of liver fibrosis.

|  | **No fibrosis**<br>**n=15** | **Early**<br>**fibrosis**<br>**n=50** | **Advance**<br>**d fibrosis**<br>**n=41** | **p value** |
| --- | --- | --- | --- | --- |
| **Left systemic ventricle**<br>**morphology, n (%)** | 8 (53.3) | 35 (70.0) | 20 (48.8) | 0.078 |
| **Age at Fontan (median,**<br>**IQR)** | 4.0 (6.0) | 5.0 (7.0) | 4.0 (5.0) | 0.722 |
| **Type of Fontan, n (%)** |  |  |  | 0.363 |
| Atriopulmonary | 3 (20.0) | 19 (38.0) | 12 (29.3) |  |
| Bjork | 0 (0.0) | 1 (2.0) | 1 (2.4) |  |
| Extracardiac | 4 (26.7) | 5 (10.0) | 11 (26.8) |  |
| Lateral tunnel (Intra-atrial<br>included here) | 8 (53.3) | 23 (46.0) | 14 (34.2) |  |
| Other | 0 (0.0) | 2 (4.0) | 3 (7.3) |  |
| **Fenestration, n %**<br>**(patients with available**<br>**data n=93)** | 5 (38.5) | 12 (27.9) | 16 (43.2) | 0.350 |
| **Fenestration closure, n %**<br>**(patients with available**<br>**data n=30)** | 3 (60.0) | 7 (63.6) | 12 (85.7) | 0.353 |
| **Fontan conversion, n %** | 2 (13.3) | 10 (20.0) | 15 (36.6) | 0.099 |
| **Heterotaxy, n %** | 1 (6.7) | 5 (10.0) | 7 (17.1) | 0.459 |
| **Diabetes, n %** | 0 (0.0) | 2 (4.0) | 4 (9.8) | 0.428 |
| **Hypertension, n %** | 1 (6.7) | 3 (6.0) | 5 (12.2) | 0.712 |
| **Hepatitis C, n %** | 0 (0.0) | 2 (4.0) | 3 (7.3) | 0.693 |
| **Obesity, n %** | 1 (6.7) | 8 (16.0) | 6 (14.6) | 0.752 |
| **BMI, median (IQR)** | 22.7 (5.7) | 23.8 (7.6) | 24.2 (7.3) | 0.957 |
| **Hemidiaphragm paresis,**<br>**n (%)** | 0 (0.0) | 1 (2.0) | 2 (4.9) | 0.735 |
| **Smoking, n (%)** | 1 (6.7) | 3 (6.0) | 3 (7.3) | 1.000 |
| **Alcohol, n (%)** | 8 (53.3) | 18 (36.0) | 13 (31.7) | 0.327 |
| **Drugs, n (%)** | 4 (26.7) | 8 (16.0) | 7 (17.1) | 0.630 |
| **Prior thromboembolism,**<br>**n (%)** | 1 (6.7) | 14 (28.0) | 10 (24.4) | 0.218 |
| **Protein losing**<br>**enteropathy, n (%)** | 0 (0.0) | 4 (8.0) | 5 (12.2) | 0.402 |
| **Arrhythmia, n (%)** | 6 (40.0) | 34 (68.0) | 25 (61.0) | 0.148 |
| **Pacemaker, n (%)** | 4 (26.7) | 19 (38.0) | 20 (48.8) | 0.286 |
| **Defibrillator, n (%)** | 1 (6.7) | 1 (2.0) | 3 (7.3) | 0.352 |
| **NYHA class, n (%)** |  |  |  | 0.064 |
| **I** | 12 (80.0) | 26 (52.0) | 15 (36.6) |  |
| **II** | 3 (20.0) | 17 (34.0) | 21 (51.2) |  |
| **III** | 0 (0.0) | 5 (10.0) | 3 (7.3) |  |
| **IV** | 0 (0.0) | 0 (0.0) | 2 (4.9) |  |
| **Betablockers, n (%)** | 8 (53.3) | 24 (48.0) | 18 (43.9) | 0.811 |
| **PDE-5 inhibitors, n (%)** | 1 (6.7) | 19 (38.0) | 18 (43.9) | **0.033** |
| **ERAs, n (%)** | 1 (6.7) | 2 (4.0) | 2 (4.9) | 0.841 |
| **Antiplatelets, n (%)** | 9 (60.0) | 25 (50.0) | 23 (56.1) | 0.737 |
| **Anticoagulants, n (%)** | 4 (26.7) | 27 (54.0) | 19 (46.3) | 0.176 |
| **Loop diuretics, n (%)** | 5 (33.3) | 28 (56.0) | 25 (61.0) | 0.178 |
| **ACE inhibitors, n (%)** | 9 (60.0) | 23 (46.0) | 19 (46.3) | 0.610 |
| **ARB, n (%)** | 1 (6.7) | 6 (12.0) | 2 (4.9) | 0.516 |
| **Aldosterone antagonists, n (%)** | 3 (20.0) | 17 (34.0) | 21 (51.2) | 0.068 |
| **Digoxin, n (%)** | 3 (20.0) | 9 (18.0) | 10 (24.4) | 0.754 |
| **Other antiarrhythmics, n (%)** | 2 (13.3) | 16 (32.0) | 8 (19.5) | 0.214 |
| **Creatinine (median, IQR)** | 0.8 (0.2) | 0.8 (0.2) | 0.9 (0.4) | 0.223 |
| **Platelets (median, IQR)** | 170.0 (53.0) | 158.0 (82.0) | 134.5 (60.0) | 0.106 |
| **Hemoglobin (median, IQR)** | 15.5 (4.0) | 15.1 (2.9) | 15.1 (2.7) | 0.692 |
| **Albumin (median, IQR)** | 4.7 (0.6) | 4.7 (0.7) | 4.4 (1.0) | 0.057 |
| **INR (median, IQR)** | 1.2 (0.0) | 1.2 (0.3) | 1.3 (0.4) | 0.230 |
| **AFP (median, IQR)** | 3.0 (8.0) | 2.0 (1.0) | 3.0 (2.0) | 0.489 |
| **BNP (median, IQR)** | 44.0 (46.0) | 47.0 (91.0) | 95.0 (130.0) | **0.016** |
| **Systemic ventricle function, n (%)** |  |  |  | 0.181 |
| Normal or mildly reduced | 12 (80.0) | 45 (90.0) | 31 (75.6) |  |
| Moderately or severely reduced | 3 (20.0) | 5 (10.0) | 10 (24.4) |  |
| **AVV regurgitation, n (%)** |  |  |  | 0.161 |
| None, trace or mild | 13 (86.7) | 36 (72.0) | 25 (61.0) |  |
| Moderate or severe | 2 (13.3) | 14 (28.0) | 16 (39.0) |  |
| **Liver US characteristics,**<br>**n (%) (patients with**<br>**available data n=88)** |  |  |  |  |
| Nodular contour | 5 (35.7) | 24 (63.2.0) | 24 (66.7) | 0.118 |
| Splenomegaly | 4 (28.6) | 11 (29.0) | 13 (36.1) | 0.772 |
| Abdominal collateral<br>vessels | 1 (7.1) | 0 (0.0) | 1 (2.8) | 0.155 |
| Ascites | 0 (0.0) | 7 (18.4) | 8 (22.2) | 0.165 |
| Elevated liver stiffness | 4 (28.6) | 14 (36.8) | 14 (38.9) | 0.790 |
| Liver mass | 0 (0.0) | 3 (7.9) | 1 (2.8) | 0.514 |
| **Liver MRI characteristics,**<br>**n (%) (patients with**<br>**available data n=25)** |  |  |  |  |
| Nodular contour | 5 (100.0) | 9 (64.3) | 4 (66.7) | 0.405 |
| Splenomegaly | 1 (20.0) | 4 (28.6) | 3 (50.0) | 0.611 |
| Ascites | 0 (0.0) | 2 (14.3) | 2 (33.3) | 0.468 |
| Abdominal collateral | 0 (0.0) | 1 (7.1) | 1 (16.7) | 0.697 |
| vessels |  |  |  |  |
| Elevated stiffness | 0 (0.0) | 0 (0.0) | 2 (33.3) | 0.083 |
| Liver mass | 0 (0.0) | 0 (0.0) | 2 (33.3) | 0.083 |
| **Liver CT characteristics,**<br>**n (%) (patients with**<br>**available data n=54)** |  |  |  |  |
| Nodular contour | 4 (66.7) | 21 (80.8) | 20 (90.9) | 0.275 |
| Splenomegaly | 4 (66.7) | 8 (30.8) | 15 (68.2) | 0.032 |
| Abdominal collateral<br>vessels | 2 (33.3) | 6 (23.1) | 10 (45.5) | 0.270 |
| Ascites | 3 (50.0) | 11 (42.3) | 11 (50.0) | 0.926 |
| Liver mass | 0 (0.0) | 3 (11.5) | 2 (9.1) | 1.000 |
| **Baseline O2Sat (median,**<br>**IQR) (patients with**<br>**available data n=92)** | 93.0 (3.0) | 92.5 (6.0) | 92.0 (6.0) | 0.655 |
| **Peak O2Sat (median, IQR)** | 92.0 (5.0) | 89.0 (8.0) | 90.0 (7.0) | 0.616 |
| **Exercise-induced**<br>**desaturation (peak O2 –**<br>**baseline O2) (median,**<br>**IQR)** | 2.0 (2.0) | 3.0 (5.0) | 2.0 (3.0) | 0.576 |
| **Peak VO2 (median, IQR)** | 61.0 (17.0) | 59.0 (22.0) | 55.5<br>(26.0) | 0.645 |
| **VE/VCO2 slope (median, IQR)** | 27.1 (6.0) | 27.7 (5.8) | 30.9 (6.4) | **0.009** |
| **High exercise capacity, n (%)** | 2 (13.3) | 6 (12.0) | 7 (17.1) | n/a |
| **Low exercise capacity, n (%)** | 2 (13.3) | 9 (18.0) | 13 (31.7) | n/a |
| **Fontan pressure, median (IQR)** | 14.0 (5.0) | 14.5 (3.0) | 17.0 (6.0) | **<.001** |
| **Mean PAP, median (IQR)** | 13.5 (5.0) | 14.0 (3.0) | 17.0 (5.0) | **<.001** |
| **Pulmonary capillary wedge pressure, median (IQR)** | 10.0 (4.0) | 10.0 (4.0) | 12.0 (5.0) | **0.005** |
| **Single ventricle end-diastolic pressure, median (IQR)** | 10.0 (5.0) | 10.0 (5.5) | 10.0 (3.0) | 0.609 |
| **Qs, median (IQR)** | 4.3 (1.4) | 4.5 (1.9) | 5.0 (2.6) | 0.273 |
| **Qs index, median (IQR)** | 2.5 (0.7) | 2.7 (1.1) | 2.7 (1.6) | 0.153 |
| **PVR, median (IQR)** | 0.9 (0.4) | 0.9 (0.7) | 1.1 (0.8) | 0.485 |
| **PVRi, median (IQR)** | 1.7 (0.8) | 1.6 (1.3) | 2.0 (1.5) | 0.674 |
| **SVR, median (IQR)** | 13.0 (4.3) | 11.3 (7.3) | 10.2 (5.6) | 0.067 |
| **SVRi, median (IQR)** | 25.7 (9.2) | 19.5 (11.3) | 18.1 (9.4) | **0.014** |
| **Fontan pathway obstruction, n (%)** | 0 (0.0) | 7 (14.0) | 8 (19.5) | 0.115 |
| **Diastolic dysfunction, n (%)** | 6 (40.0) | 24 (48.0) | 29 (70.7) | **0.017** |
| **Elevated PVR, n (%)** | 4 (26.7) | 18 (36.0) | 17 (41.5) | 0.290 |
| **Aortopulmonary collaterals, n (%)** | 7 (46.7) | 5 (10.0) | 11 (26.8) | **0.006** |
| **Venous-venous collaterals, n (%)** | 8 (53.3) | 26 (52.0) | 18 (43.9) | 0.698 |

\*p-values from chi-square test for independence (or Fisher's exact test when
appropriate) for categorical variables, and Kruskal-Wallis test for continuous variables
\*\*p-values do not include missing values on any given variable
\*\*\* frequencies were calculated among those with available data and not among total N
in each column

### Composite clinical outcome and associated factors

Outcomes among patients with the Fontan circulation according to degree of liver fibrosis are found in Table 2. Those with advanced liver fibrosis on biopsy were more likely to experience the composite clinical outcome [0 (0%) without fibrosis, 8 (16.0 %) with early fibrosis and 15 (36.6%) with advanced fibrosis, p=0.005] (Figure 1). None of the individual components of the composite outcome was significantly associated with advanced fibrosis. In a multivariable Cox proportional hazards model (Table 3) controlling for age at Fontan, age at liver biopsy, right ventricular morphology, Fontan pressure, pulmonary capillary wedge pressure, VE/VCO2 slope, diastolic dysfunction, and aortopulmonary collaterals, there was no difference in the risk of the composite clinical outcome between advanced or early fibrosis. In this multivariable model, there were differences in the risk of the composite clinical outcome based on age at Fontan [HR=1.42 (SE 0.18); p=0.045], right ventricular morphology [HR=26.01 (SE 1.53); p=0.033], and presence of aortopulmonary collaterals [HR=122.77 (SE 1.63); p=0.003].

**Figure 1.**
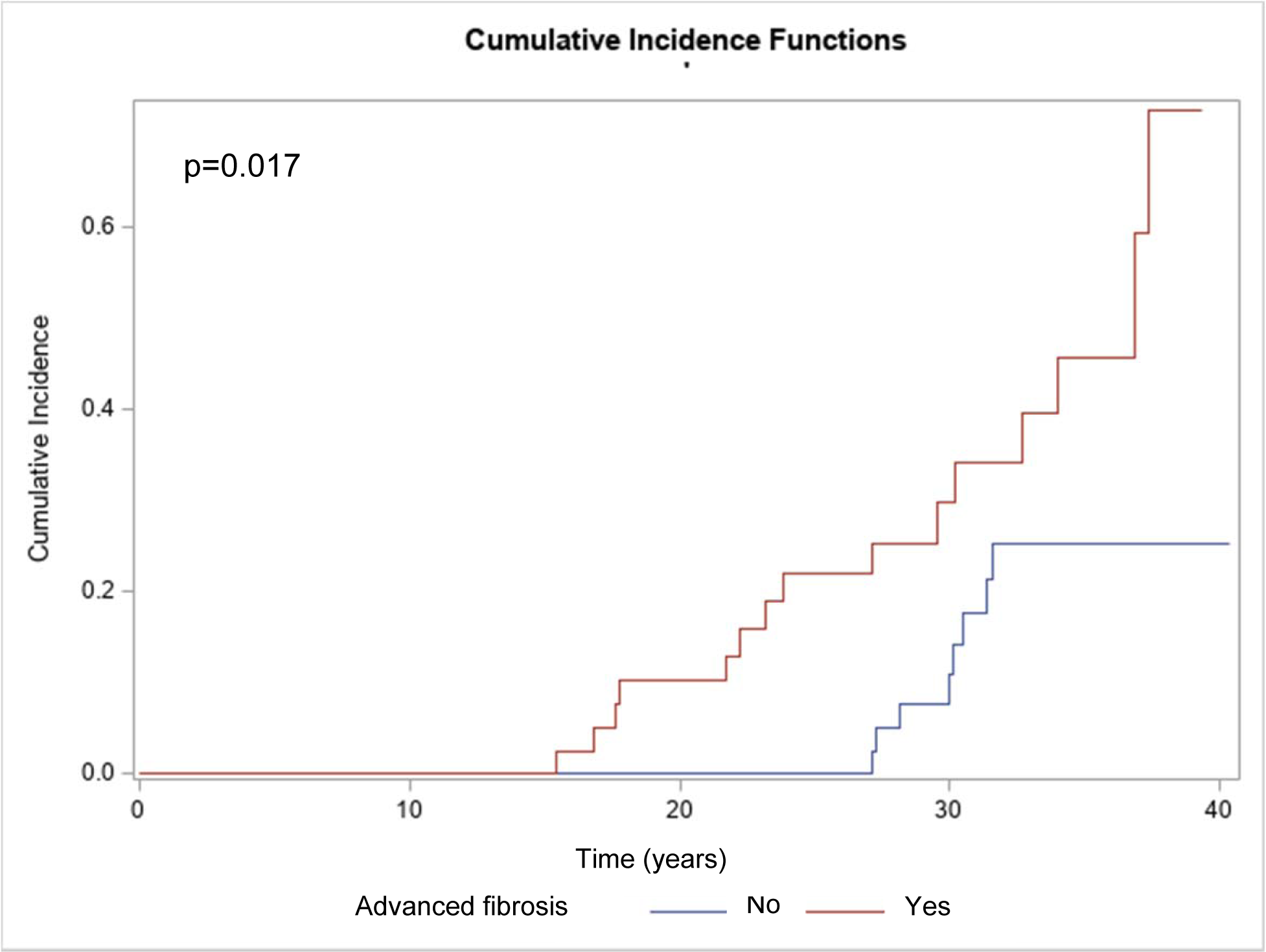
Unadjusted cumulative incidence function comparing time to the clinical composite outcome, relative to date of Fontan, between patients with early vs advanced fibrosis.

**Table 2.** Outcomes among patients with the Fontan circulation according to liver biopsy results.

| **Outcomes** | **No fibrosis** | **Early**<br>**fibrosis** | **Advanced**<br>**fibrosis** | **P value** |
| --- | --- | --- | --- | --- |
| **Thromboembolism,**<br>**n (%)** | 1 (6.7) | 1 (2.0) | 1 (2.4) | 0.527 |
| **Arrhythmias, n (%)** | 4 (26.7) | 14 (28.0) | 7 (17.0) | 0.215 |
| **Composite clinical**<br>**outcome, n (%)** | 0 (0) | 8 (16.0) | 15 (36.6) | **0.005** |
| **Decompensated**<br>**cirrhosis, n (%)** | 0 (0) | 4 (8.0) | 7 (17.1) | 0.138 |
| Ascites requiring<br>paracentesis | 0 (0) | 3 (6.0) | 5 (12.2) | 0.402 |
| Esophageal variceal<br>bleeding | 0 (0) | 1 (2.0) | 2 (4.9) | 0.740 |
| Hepatic<br>encephalopathy | 0 (0) | 0 (0.0) | 0 (0.0) | n/a |
| **Hepatocellular**<br>**carcinoma, n (%)** | 0 (0.0) | 2 (4.0) | 1 (2.4) | 1.000 |
| **Focal nodular** | 1 (6.7) | 5 (10.0) | 5 (12.2) | 0.544 |
| **hyperplasia, n (%)** |  |  |  |  |
| **Chronic kidney disease, n (%)** | 0 (0.0) | 1 (2.0) | 1 (2.4) | 0.115 |
| **Fontan circulatory failure, n (%)** | 1 (6.7) | 11 (22.0) | 15 (36.6) | 0.056 |
| **Liver transplant, n (%)** | 0 (0.0) | 5 (10.0) | 8 (19.5) | 0.109 |
| **Heart transplant, n (%)** | 1 (6.7) | 9 (18.0) | 13 (31.7) | 0.090 |
| **Death, n (%)** | 0 (0.0) | 3 (6.0) | 3 (7.3) | 0.858 |

\*Composite clinical outcome of decompensated cirrhosis (ascites requiring paracentesis, esophageal variceal bleeding, hepatic encephalopathy), hepatocellular carcinoma, need for liver transplantation, protein-losing enteropathy, chronic kidney disease stage 4 or higher, or death.

**Table 3.** Multivariable Cox proportional hazards model for the composite clinical outcome.

| Parameter | Hazard Ratio | Standard Error | P value |
| --- | --- | --- | --- |
| Age at Fontan | 1.42 | 0.18 | **0.045** |
| Right ventricular morphology | 26.01 | 1.53 | **0.033** |
| Fontan pressure | 1.68 | 0.37 | 0.162 |
| Pulmonary capillary wedge pressure | 1.17 | 0.24 | 0.517 |
| VE/VCO2 slope | 0.97 | 0.09 | 0.744 |
| Diastolic dysfunction | 0.08 | 2.00 | 0.209 |
| Aortopulmonary collaterals | 122.77 | 1.62 | **0.003** |
| Age at liver biopsy | 0.97 | 0.10 | 0.752 |
| Advanced fibrosis | 0.19 | 1.22 | 0.179 |

### FALD has distinct mRNA expression profiles according to degree of fibrosis and composite clinical outcome

Liver samples from patients with advanced fibrosis had 231 DEGs (228 up-regulated) compared to those with early fibrosis (Figure 2A). Liver samples from patients with the composite clinical outcome had 906 DEGs (894 up-regulated) compared to those without it (Figure 2B). A total of 136 DEGs were identified in both comparisons and they were enriched in various cellular responses and signaling pathways, including response to wounding, extracellular matrix organization, regulation of cell adhesion, IL-18, MAPK and TGF-beta signaling pathways, vasculature development and angiogenesis (Figure 3A). Significant correlations of these 136 DEGs were found with various diseases from DisGeNET database, including idiopathic pulmonary arterial hypertension, lung diseases, cardiac fibrosis, vascular diseases, and endothelial dysfunction (Figure 3B). The distribution of specific DEGs involved in FALD pathophysiology are shown according to the degree of fibrosis (Figure 4) and presence or absence of the clinical outcome (Figure 5).

**Figure 2.**
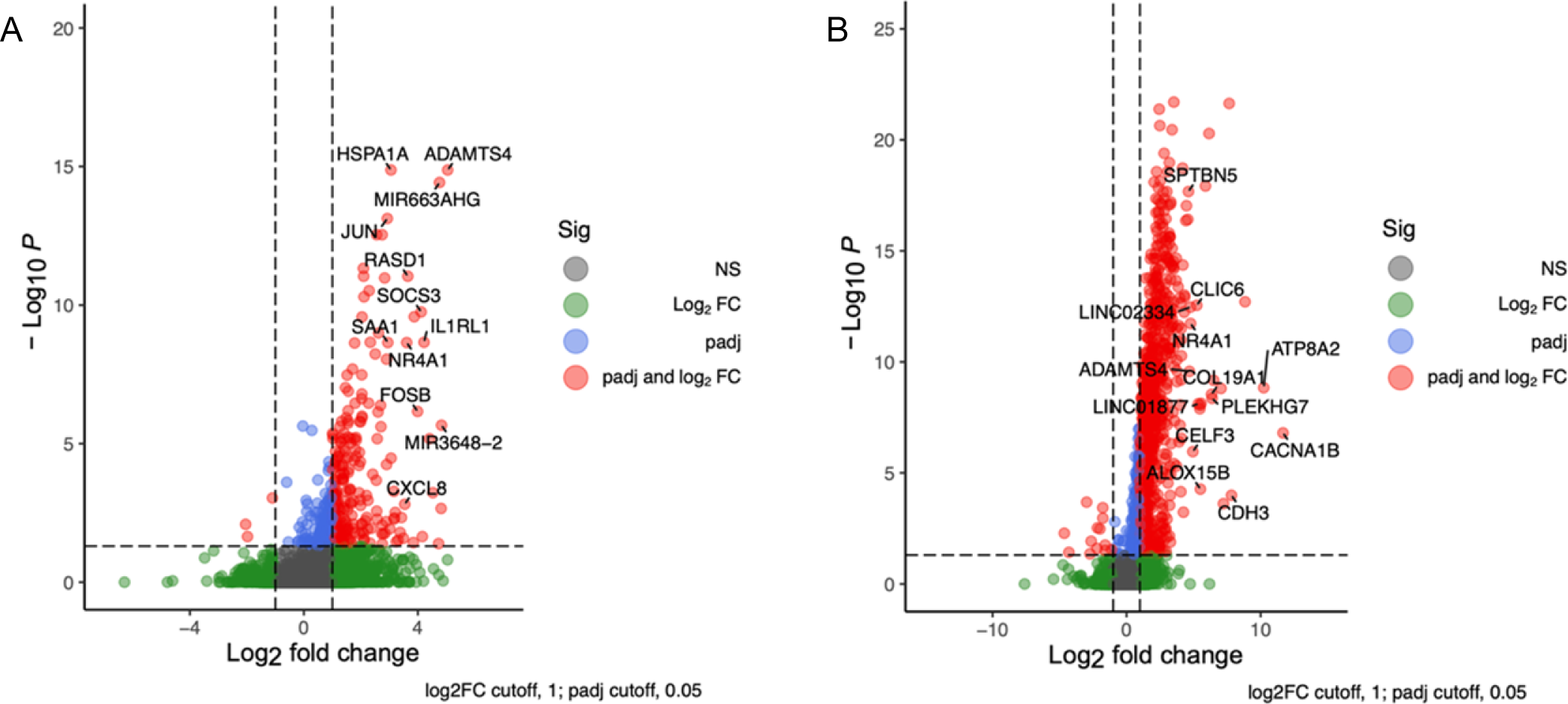
A. Volcano plot showing 231 DEGs (228 up-regulated) in patients with FALD and advanced fibrosis vs early fibrosis. B. Volcano plot showing 906 DEGs (894 up-regulated) in patients with FALD and the clinical composite outcomes vs those without it.

**Figure 3.**
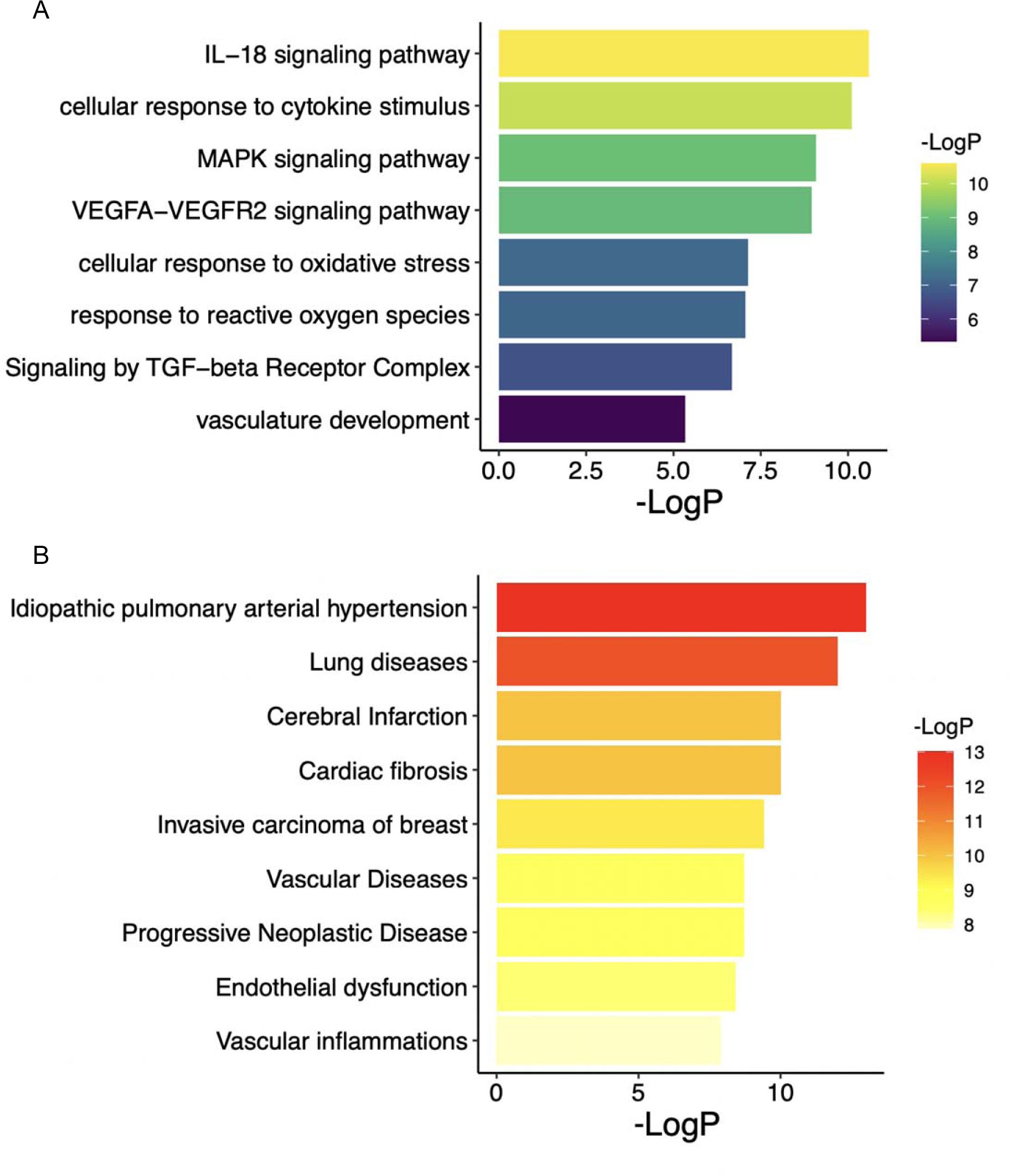
A. Gene ontology and pathway enrichment analysis among patients with advanced liver fibrosis. B. Gene disease association enrichment analysis using 136 DEGs and the DisGeNet platform.

**Figure 4.**
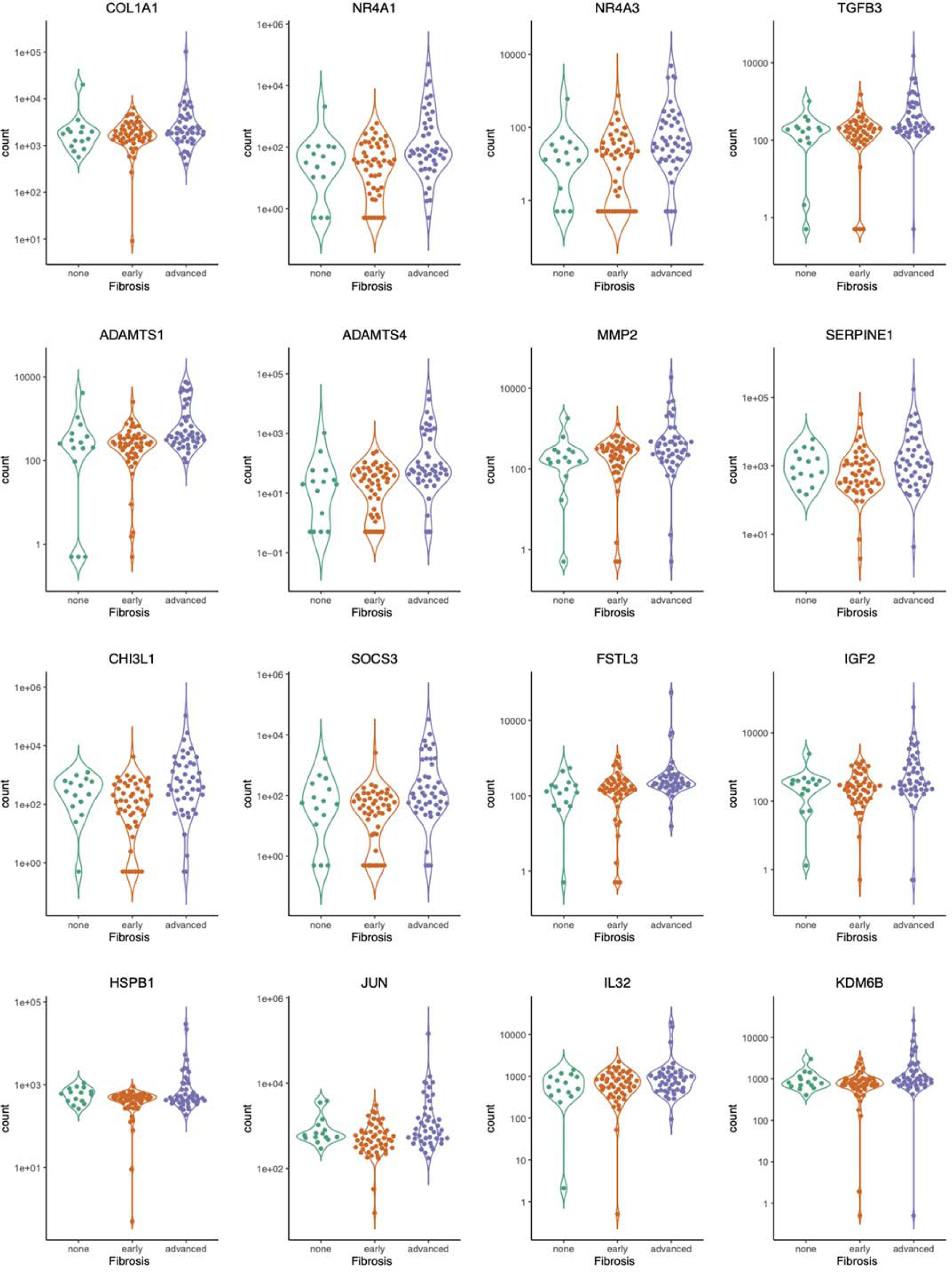
Violin plot demonstrating the gene count distribution in specific overlapping genes among patients with FALD according to the degree of liver fibrosis.

**Figure 5.**
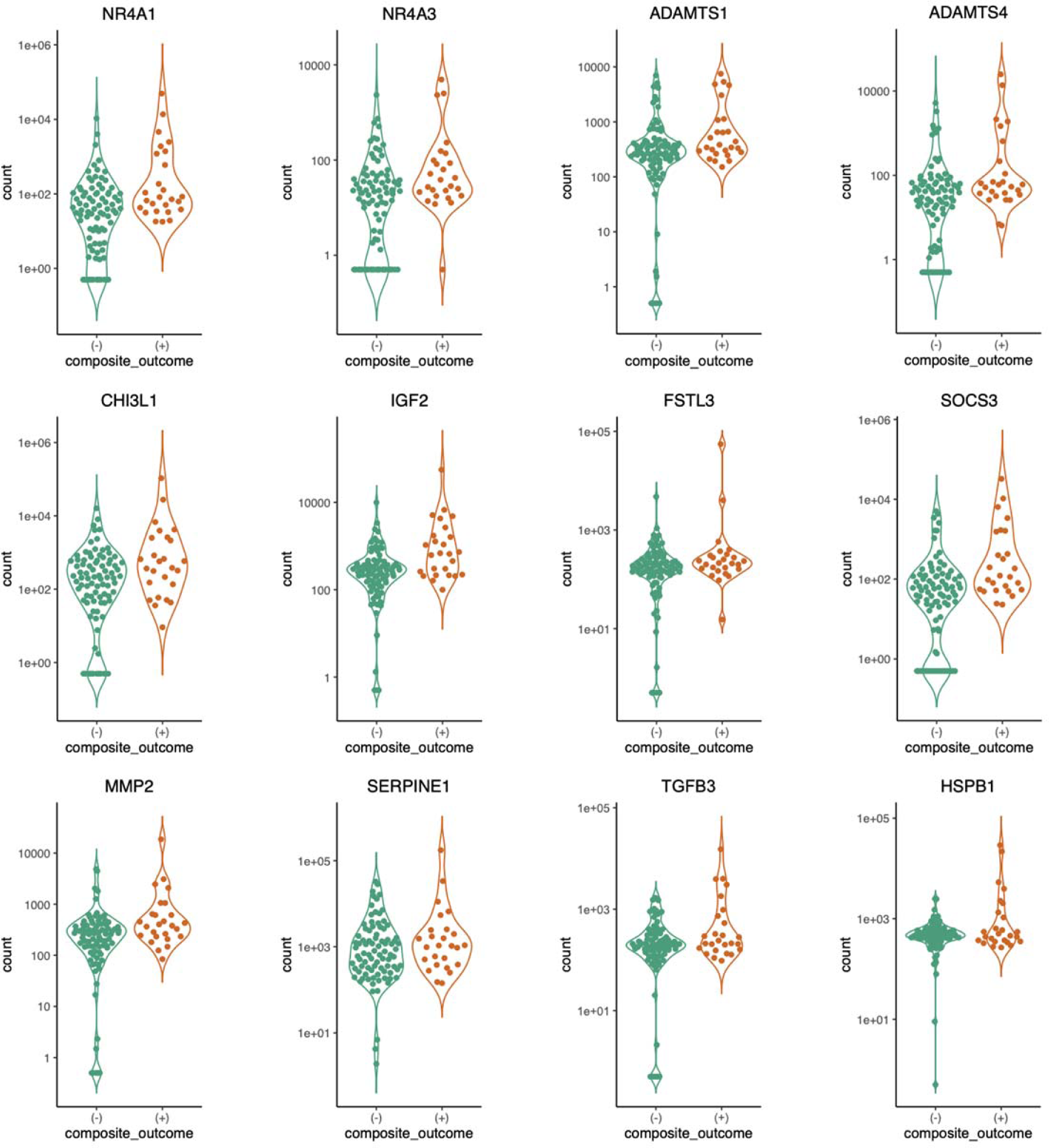
Violin plot demonstrating the gene count distribution in specific overlapping genes among patients with FALD according to the presence of the composite outcome.

## DISCUSSION

In this first broad analysis of gene expression in liver tissue from a retrospectively identified cohort of patients with FALD, we reveal several important findings. First, patients with FALD and advanced fibrosis have a distinct transcriptome versus those with early fibrosis and controls. Second, patients with FALD and advanced fibrosis are more likely to experience the composite clinical outcome, but this is mediated by age at Fontan, right ventricular morphology and presence of aortopulmonary collaterals. Third, patients with FALD and the composite clinical outcome have a distinct transcriptome compared to those without the composite clinical outcome. Fourth, we identified overlapping DEGs between those with advanced fibrosis as well as the composite clinical outcome, and identified pathways related to pro-inflammatory responses and increased oxidative stress (HSPB1, IRF1, SOCS3, and NFKB2), impaired vascular endothelial function (HSPA1A, HSPA1B, ND5, ND6, NR4A2, SOD3, KLF2), enriched angiogenesis and vasculature development (RHOB, COL1A1, NR4A1, NOTCH3, SERPINE1), TGF-beta signaling pathways, etc. These findings are important given that an inflammatory infiltrate on liver biopsies was not found, and we excluded samples with active hepatocellular carcinoma. These results expand our insights into FALD pathophysiology and show potential candidate genes that could serve as biomarkers of adverse outcomes.

Out of all the overlapping DEGs identified in our study, COL1A1 RASD1, CHI3L1, MTRNR2, MUC5B, SLCO4A1, SOD3, WNK2 have been previously described in transcriptomic profiling of obesity-related nonalcoholic steatohepatitis^15^. Our findings demonstrate that angiogenesis may be particularly important in FALD. This complex, dynamic and growth factor dependent process leading to the formation of new blood vessels from preexisting ones, is strongly associated with scar formation and sinusoidal remodeling in chronic liver diseases^10^. Several genes are involved in this process including VEGF and ANGPT2^16^. High serum angiopoietin-2 levels (protein expressed from ANGPT2) have been associated with liver cirrhosis and hepatocellular carcinoma among samples from nonalcoholic steatohepatitis^17^, and several studies are evaluating this as a potential therapeutic target^18–20^. In a cohort study involving patients with the Fontan circulation, angiopoietin-2 levels were found to be significantly higher in patients with active or recent arrhythmias^21^. Although we did not find ANGPT2 as one of our overlapping genes, we identified VEGFA-VEGFR2 signaling pathway, which is the major pathway that activates angiogenesis.

Our study had several strengths, including a highly diverse and phenotypically characterized cohort that had longitudinal care at a large ACHD referral center with expertise in combined heart and liver transplantation, as well as achieving more than 80% high-quality RNA extraction from FFPE liver biopsies. Moreover, the patients had undergone cardiac catheterization in a standard fashion, limiting the heterogeneity of hemodynamic measurements. Despite this, several limitations were encountered. First, intrahepatic gene expression profiles are largely driven by the effect of parenchymal cells, and this represents a limitation when analyzing bulk transcriptomics. Several recent discoveries in the molecular biology of liver cirrhosis (from non-cardiac causes) have highlighted the important role that non-parenchymal liver cells (immune, endothelial, and mesenchymal cells) play in its development. Gene expression profiles of non-parenchymal cells are underrepresented in the whole tissue liver RNA-seq data. Other methods such as single-cell RNA sequencing or digital spatial profiling^22^ may be helpful identifying the gatekeepers of advanced liver fibrosis in FALD. Second, the assessment of aortopulmonary collaterals was not systematically performed in all patients unless the patients presented for organ transplantation, and this might influence our univariate as well as multivariable Cox proportional hazards results. Third, small cohort size could have influenced the significance of our results.

## CONCLUSIONS

Patients with Fontan-associated liver disease exhibit transcriptomic differences according to the degree of fibrosis and the presence of the clinical composite outcome. These genes are involved in pathways related to inflammation, congestion, and angiogenesis.

## Data Availability

The data can be accessed upon reasonable request.

## SUPPLEMENTAL FIGURES

**Supplemental figure 1.**
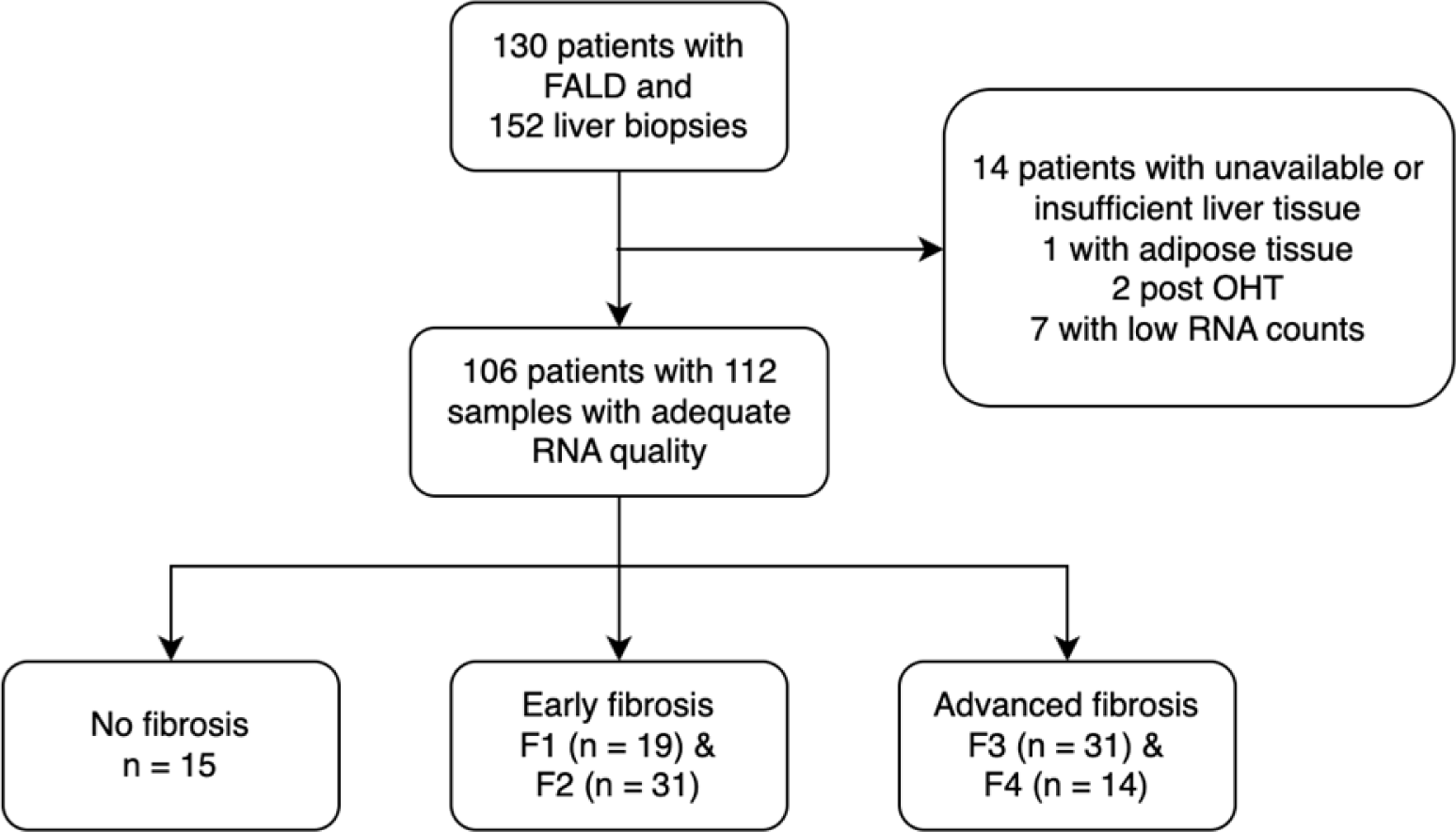
Flow diagram of patients and samples included.

## REFERENCES

1. Fontan F, Baudet E. Surgical repair of tricuspid atresia. Thorax. 1971;26:240–248. doi: 10.1136/thx.26.3.240

2. Emamaullee J, Zaidi AN, Schiano T, Kahn J, Valentino PL, Hofer RE, Taner T, Wald JW, Olthoff KM, Bucuvalas J, et al. Fontan-Associated Liver Disease: Screening, Management, and Transplant Considerations. Circulation. 2020;142:591–604. doi: 10.1161/CIRCULATIONAHA.120.045597

3. Agnoletti G, Ferraro G, Bordese R, Marini D, Gala S, Bergamasco L, Ferroni F, Calvo PL, Barletti C, Cisaro F, et al. Fontan circulation causes early, severe liver damage. Should we offer patients a tailored strategy? Int J Cardiol. 2016;209:60–65. doi: 10.1016/j.ijcard.2016.02.041

4. Goldberg DJ, Surrey LF, Glatz AC, Dodds K, O’Byrne ML, Lin HC, Fogel M, Rome JJ, Rand EB, Russo P, et al. Hepatic Fibrosis Is Universal Following Fontan Operation, and Severity is Associated With Time From Surgery: A Liver Biopsy and Hemodynamic Study. J Am Heart Assoc. 2017;6. doi: 10.1161/JAHA.116.004809

5. Khan S, Aziz H, Emamaullee J. Research priorities in Fontan-associated liver disease. Curr Opin Organ Transplant. 2020;25:489–495. doi: 10.1097/MOT.0000000000000803

6. Egbe AC, Poterucha JT, Warnes CA, Connolly HM, Baskar S, Ginde S, Clift P, Kogon B, Book WM, Walker N, et al. Hepatocellular Carcinoma After Fontan Operation: Multicenter Case Series. Circulation. 2018;138:746–748. doi: 10.1161/CIRCULATIONAHA.117.032717

7. Rychik J, Veldtman G, Rand E, Russo P, Rome JJ, Krok K, Goldberg DJ, Cahill AM, Wells RG. The precarious state of the liver after a Fontan operation: summary of a multidisciplinary symposium. Pediatr Cardiol. 2012;33:1001–1012. doi: 10.1007/s00246-012-0315-7

8. Keung CY, Zentner D, Gibson RN, Phan DH, Grigg LE, Sood S, Nicoll AJ. Fontan-associated liver disease: pathophysiology, investigations, predictors of severity and management. Eur J Gastroenterol Hepatol. 2019. doi: 10.1097/MEG.0000000000001641

9. Hilscher MB, Johnson JN, Cetta F, Driscoll DJ, Poterucha JJ, Sanchez W, Connolly HM, Kamath PS. Surveillance for liver complications after the Fontan procedure. Congenit Heart Dis. 2017;12:124–132. doi: 10.1111/chd.12446

10. Alsaied T, Rathod RH, Aboulhosn JA, Budts W, Anderson JB, Baumgartner H, Brown DW, Cordina R, D’Udekem Y, Ginde S, et al. Reaching consensus for unified medical language in Fontan care. ESC Heart Fail. 2021;8:3894–3905. doi: 10.1002/ehf2.13294

11. Dobin A, Davis CA, Schlesinger F, Drenkow J, Zaleski C, Jha S, Batut P, Chaisson M, Gingeras TR. STAR: ultrafast universal RNA-seq aligner. Bioinformatics. 2013;29:15–21. doi: 10.1093/bioinformatics/bts635

12. Zhou Y, Zhou B, Pache L, Chang M, Khodabakhshi AH, Tanaseichuk O, Benner C, Chanda SK. Metascape provides a biologist-oriented resource for the analysis of systems-level datasets. Nat Commun. 2019;10:1523. doi: 10.1038/s41467-019-09234-6

13. Pinero J, Bravo A, Queralt-Rosinach N, Gutierrez-Sacristan A, Deu-Pons J, Centeno E, Garcia-Garcia J, Sanz F, Furlong LI. DisGeNET: a comprehensive platform integrating information on human disease-associated genes and variants. Nucleic Acids Res. 2017;45:D833–D839. doi: 10.1093/nar/gkw943

14. Pinero J, Queralt-Rosinach N, Bravo A, Deu-Pons J, Bauer-Mehren A, Baron M, Sanz F, Furlong LI. DisGeNET: a discovery platform for the dynamical exploration of human diseases and their genes. Database (Oxford). 2015;2015:bav028. doi: 10.1093/database/bav028

15. Gerhard GS, Legendre C, Still CD, Chu X, Petrick A, DiStefano JK. Transcriptomic Profiling of Obesity-Related Nonalcoholic Steatohepatitis Reveals a Core Set of Fibrosis-Specific Genes. J Endocr Soc. 2018;2:710–726. doi: 10.1210/js.2018-00122

16. Bocca C, Novo E, Miglietta A, Parola M. Angiogenesis and Fibrogenesis in Chronic Liver Diseases. Cell Mol Gastroenterol Hepatol. 2015;1:477–488. doi: 10.1016/j.jcmgh.2015.06.011

17. Lefere S, Devisscher L, Geerts A. Angiogenesis in the progression of non-alcoholic fatty liver disease. Acta Gastroenterol Belg. 2020;83:301–307.

18. Scholz A, Rehm VA, Rieke S, Derkow K, Schulz P, Neumann K, Koch I, Pascu M, Wiedenmann B, Berg T, et al. Angiopoietin-2 serum levels are elevated in patients with liver cirrhosis and hepatocellular carcinoma. Am J Gastroenterol. 2007;102:2471–2481. doi: 10.1111/j.1572-0241.2007.01377.x

19. Pauta M, Ribera J, Melgar-Lesmes P, Casals G, Rodriguez-Vita J, Reichenbach V, Fernandez-Varo G, Morales-Romero B, Bataller R, Michelena J, et al. Overexpression of angiopoietin-2 in rats and patients with liver fibrosis. Therapeutic consequences of its inhibition. Liver Int. 2015;35:1383–1392. doi: 10.1111/liv.12505

20. Lefere S, Van de Velde F, Hoorens A, Raevens S, Van Campenhout S, Vandierendonck A, Neyt S, Vandeghinste B, Vanhove C, Debbaut C, et al. Angiopoietin-2 Promotes Pathological Angiogenesis and Is a Therapeutic Target in Murine Nonalcoholic Fatty Liver Disease. Hepatology. 2019;69:1087–1104. doi: 10.1002/hep.30294

21. Shirali AS, Lluri G, Guihard PJ, Conrad MB, Kim H, Pawlikowska L, Bostrom KI, Iruela-Arispe ML, Aboulhosn JA. Angiopoietin-2 predicts morbidity in adults with Fontan physiology. Sci Rep. 2019;9:18328. doi: 10.1038/s41598-019-54776-w

22. Lee J, Kim CM, Cha JH, Park JY, Yu YS, Wang HJ, Sung PS, Jung ES, Bae SH. Multiplexed Digital Spatial Protein Profiling Reveals Distinct Phenotypes of Mononuclear Phagocytes in Livers with Advanced Fibrosis. Cells. 2022;11. doi: 10.3390/cells11213387

